# The *NR5A1/SF-1* variant p.Gly146Ala cannot explain the phenotype of individuals with a difference of sex development

**DOI:** 10.1101/2023.02.13.23285760

**Authors:** Idoia Martinez de Lapiscina, Chrysanthi Kouri, Josu Aurrekoetxea, Mirian Sanchez, Rawda Naamneh Elzenaty, Kay-Sara Sauter, Núria Camats, Gema Grau, Itxaso Rica, Amaia Rodriguez, Amaia Vela, Alicia Cortazar, M. Concepción Alonso-Cerezo, Pilar Bahillo, Laura Berthod, Isabel Esteva, Luis Castaño, Christa E. Flück

## Abstract

Steroidogenic factor 1 (SF-1, *NR5A1*) plays an important role in human sex development. Variants of *NR5A1/*SF-1 may cause mild to severe differences of sex development (DSD) or may be found in healthy carriers. So far, the broad DSD phenotypic variability associated *NR5A1*/SF-1 variants remains a conundrum. The *NR5A1*/SF-1 variant c.437G>C/p.Gly146Ala is common in individuals with a DSD and has been suggested to act as a susceptibility factor for adrenal disease or cryptorchidism. However, as the allele frequency in the general population is high, and as functional testing of the p.Gly146Ala variant *in vitro* revealed inconclusive results, the disease-causing effect of this variant has been questioned. However, a role as a disease modifier in concert with other gene variants is still possible given that oligogenic inheritance has been described in patients with *NR5A1*/SF-1 gene variants. Therefore, we performed next generation sequencing in DSD individuals harboring the *NR5A1*/SF-1 p.Gly146Ala variant to search for other DSD-causing variants. Aim was to clarify the function of this variant for the phenotype of the carriers. We studied 14 pediatric DSD individuals who carried the p.Gly146Ala variant. Panel and whole-exome sequencing was performed, and data were analyzed with a specific data filtering algorithm for detecting variants in *NR5A1*- and DSD-related genes. The phenotype of the studied individuals ranged from scrotal hypospadias and ambiguous genitalia in 46,XY DSD to typical male external genitalia and ovotestes in 46,XX DSD patients. Patients were of African, Spanish, and Asian origin. Of the 14 studied subjects, five were homozygous and nine heterozygous for the *NR5A1*/SF-1 p.Gly146Ala variant. In ten subjects we identified either a clearly pathogenic DSD gene variant (e.g. in *AR, LHCGR*) or one to four potentially deleterious variants that likely explain the observed phenotype alone (e.g. in *FGFR3, CHD7, ADAMTS16*). Our study shows that most individuals carrying the *NR5A1*/SF-1 p.Gly146Ala variant, harbor at least one other deleterious gene variant which can explain the DSD phenotype. This finding confirms that the p.Gly146Ala variant of *NR5A1/SF-1* may not contribute to the pathogenesis of DSD and qualifies as a benign polymorphism. Thus, individuals, in whom the *NR5A1*/SF-1 p.Gly146Ala gene variant has been identified as the underlying genetic cause for their DSD in the past, should be re-evaluated with a next-generation sequencing method to reveal the real genetic diagnosis.

## Introduction

Typical sex development depends on the specific interactions of many genes and pathways in a defined spatiotemporal sequence. Any perturbation in this very complex biological event, which involves genetic and hormonal processes, may result in atypical sex development and leads to a broad range of differences of sex development (DSD) in humans, also known as variations of sex characteristics (VSC) (1). The *NR5A1* gene, which encodes the steroidogenic factor 1 (SF-1), regulates multiple genes implicated in adrenal development, gonadal determination and differentiation, steroidogenesis, and reproduction (2). Since the identification of the first *NR5A1*/SF-1 variation in a 46,XY patient with primary adrenal failure and complete gonadal dysgenesis (3), the gonadal and reproductive phenotype associated with *NR5A1*/SF-1 variants has broadened including 46,XY and 46,XX cases with sex reversal to minor untypicalities such as hypospadias or even healthy carriers, whereas adrenal failure has only been found in very rare cases (4, 5).

Currently, 291 different *NR5A1*/SF-1 variants are reported in the Human Gene Mutation Database (HGMD, October 2022). Variants include missense/nonsense, indels, small insertions/deletions, complete gene deletions or splice-site variants that are scattered throughout the whole gene without obvious hot spots. According to ACMG (American College of Medical Genetics and Genomics) classification (6), described variants may qualify as (likely) pathogenic or (likely) benign, and several are variants of unknown significance (VUS).

The nonsynonymous *NR5A1*/SF-1 c.437G>C/p.Gly146Ala (rs1110061) variant has been first described in a group of Japanese patients presenting with a series of adrenal diseases such as Cushing’s syndrome or non-functioning adrenocortical adenoma (7). In this context, WuQiang et al. reported that the p.Gly146Ala variant slightly impairs the transactivation ability of adrenal and ovary specific gene promoters but does not affect cofactor interaction and cellular localization (7). Later, it has been proposed to act as a susceptibility factor for cryptorchidism (8), isolated micropenis (9, 10), spermatogenic failure (11, 12), primary ovarian insufficiency (POI) (13) and type 2 diabetes (14). The p.Gly146Ala variant is common among individuals with a 46,XY DSD with a prevalence varying between 6.8 and 31% (15, 16). However, the minor allele frequency (MAF) (C allele) is also high in the overall control population (23.5%, gnomAD v3.1.2). Specifically, its MAF is increased approximately by 1.3-3-fold in the East Asian and the African control populations, whereas it is only present in 1% of the European control population (gnomAD v3.1.2). Moreover, *in vitro* studies of transcriptional activity of the *NR5A1*/SF-1 p.Gly146Ala variant on several target promoters in various cell models found unaltered wild-type functionality (15, 17). In fact, some patients who carry severe, pathogenic *NR5A1*/SF-1 variants in compound heterozygous state with the p.Gly146Ala variant, do not phenotypically differ from individuals carrying the severe variant only (5, 16-22). The DSD causing effect of the *NR5A1*/SF-1 p.Gly146Ala variant is therefore in doubt. However, given that oligogenic inheritance has been suggested for explaining the broad phenotype observed in individuals and families with *NR5A1*/SF-1 gene variants (23-29), the p.Gly146Ala variant might play a role as co-regulator or disease modifier of sexual development.

The aim of this study was therefore, to elucidate the role of the *NR5A1/*SF-1 p.Gly146Ala variant on sexual development. We studied 14 DSD patients carrying this variant by next generation sequencing (NGS). Specifically, we searched for other DSD-causing variants and their pathogenicity in order to assess the effect of the *NR5A1/*SF-1 p.Gly146Ala variant on the phenotype of its carriers.

## Patients and Methods

### Patients and ethical approval

We recruited 14 pediatric DSD individuals carrying the *NR5A1/*SF-1 p.Gly146Ala variant from a larger cohort of 125 DSD patients collected at the Biocruces Bizkaia Health Research Institute (Barakaldo, Spain). Clinical data were provided by the caring clinicians (Table 1 and S2 table). The study was approved by the ethics committee for clinical research of Euskadi (CEIC-E), Spain. Written informed consent was provided by the parents of the studied children.

**Table 1.** Phenotype of the DSD patients harbouring the *NR5A1*/SF-1 p.Gly146Ala variant. Further details including biochemical data are given in Supplementary table 2.

| Patient/<br>Origin<sup>a</sup> | Karyotype/Sex<br>assignment<sup>b</sup> | Zygoty<sup>c</sup> | Age range<br>at<br>diagnosis | External genital<br>phenotype<sup>d</sup> | Internal phenotype | Gonadal/reproductive function | Other organ<br>anomalies |
| --- | --- | --- | --- | --- | --- | --- | --- |
| 1<sup>1</sup> | 46,XY/M | Het | 0-5y | Mild | Laparoscopy: absence of female organs and gonadal tissue. Histology: undifferentiated tissue. | 0-5y, abnormal (no T response to hCG) | No |
| 2<sup>2</sup> | 46,XX/M | Het | 6-10y | Opposite sex | US: no Müllerian ducts. Histology: infantile ovary with follicular cysts, fallopian tube, atrophic uterus, mesonephric remnant. | hCG test with hyperandrogenic reaction for typical male | No |
| 3<sup>2</sup> | 46,XY/M | Het | 6-10y | Severe | US: inguinal testes (right 0.5cc; left 0.6cc). | 6-10y, normal hCG test | No |
| 4<sup>2</sup> | 46,XX/M | Hom | 6-10y | Opposite sex | Laparoscopy: bilateral gonads in inguinal canal and iliac area, atrophic uterus. No Müllerian ducts. Histology: bilateral ovotestes. | hCG test with hyperandrogenic reaction for typical male | No |
| 5<sup>2</sup> | 46,XX/M | Hom | 0-5y | Opposite sex | US: inguinal bilateral gonads (1ml), no Müllerian ducts. Histology: ovarian tissue (left), testicular and ovarian tissue (right). | hCG test with hyperandrogenic reaction for typical male | No |
| 6<sup>2</sup> | 46,XY/M | Het | 0-5y | Mild | MRI: absence of uterus and ovaries. | 0-5y, normal baseline and hCG test | Anal agenesis, iron deficiency |
| 7<sup>2</sup> | 46,XY/M | Hom | 6-10y | Severe | US: scrotal right testis (15x9mm), inguinal left testis (13x6mm). | 6-10y, normal baseline and hCG test | No |
| 8<sup>2</sup> | 46,XY/F | Het | 6-10y | Opposite sex | US: vaginal opening, no uterus. Histology: normal testis tissue at age ranges 6-10y and 21-25y. | 31-35y, abnormal (high gonadotropins and low T) | No |
| 9<sup>3</sup> | 46,XY/M | Hom | 6-10y | Typical | ND | 6-10y, high T at baseline | Pituitary adenoma, hearing loss |
| 10<sup>2</sup> | 46,XX/M | Hom | 0-5y | Opposite sex | US: prepubertal uterus. Histology: ovarian tissue (left), testicular and ovarian tissue (right). | Prepubertal | No |
| 11<sup>1</sup> | 46,XY/M | Het | 0-5y | Mild | US: normal size intrascrotal testes. | 11-15y, normal (normal T and gonadotropins) | No |
| 12<sup>3</sup> | 46,XY/M | Het | 0-5y | Severe | ND | ND | Left renal agenesis, lipomeningocele |
| 13<sup>1</sup> | 46,XY/F | Het | 11-15y | Opposite sex | Infantile uterus and ovaries by ultrasound. | ND | Abdominal obesity |
| 14<sup>1</sup> | 46,XY/M | Het | 0-5y | Severe | US: normal uterus, absence of gonads. | Abnormal (high gonadotropins and normal T) | No |

F, female; hCG, human chorionic gonadotropin; M, male; MRI, magnetic resonance imaging; ND, not determined; T, testosterone; US, ultrasound; y, years. <sup>a</sup>Origin of the patients: <sup>1</sup>Spanish; <sup>2</sup>African; <sup>3</sup>Asian. <sup>b</sup>None of the patients was sex re-assigned. <sup>c</sup>Zygoty of the *NR5A1*/SF-1 p.Gly146Ala variant identified by targeted gene panel. <sup>d</sup>Severity of the atypical phenotype of the external genitalia was classified according to (74).

Ten 46,XY DSD and four 46,XX DSD patients carrying the p.Gly146Ala variant in the *NR5A1*/SF-1 gene were analyzed using whole exome sequencing (WES) or a targeted gene panel comprised of 48 genes associated with sex determination, sex differentiation and hypogonadism (Supplementary table 1).

### Genetic analysis

Genomic DNA from the patients was extracted from peripheral blood leukocytes using the MagPurix 12S system (Zinexts Life Science Corp.). DNA purity and concentration were determined using a Qubit 2.0 fluorometer (Thermo Fisher Scientific).

Initial characterization for the *NR5A1/*SF-1 p.Gly146Ala variant was done by a DSD specific panel. Additional NGS was performed by WES (Figure 1A). For WES, libraries were prepared using the Illumina DNA/RNA Prep Tagmentation PCR reagents (Illumina) and Illumina Exome Panel (CEX) according to the manufacturer’s instructions. The resulting libraries were subjected to paired-end sequencing on a NovaSeq 6000 System (Illumina). Raw data were then converted to FastQ files for computational analysis, which included read alignment to the human genome reference sequence (GRCh38), duplicate marking, base quality score recalibration, indel realignment, and variant calling with an in-house bioinformatics pipeline using BWA-MEM (30) and GATK (31) software. Variants were annotated with wANNOVAR (32) and filtration process of the exonic variants was performed using R software (R 4.2.0). Variant filtration was performed following specific steps as given in Figure 1B.

**Figure 1.**
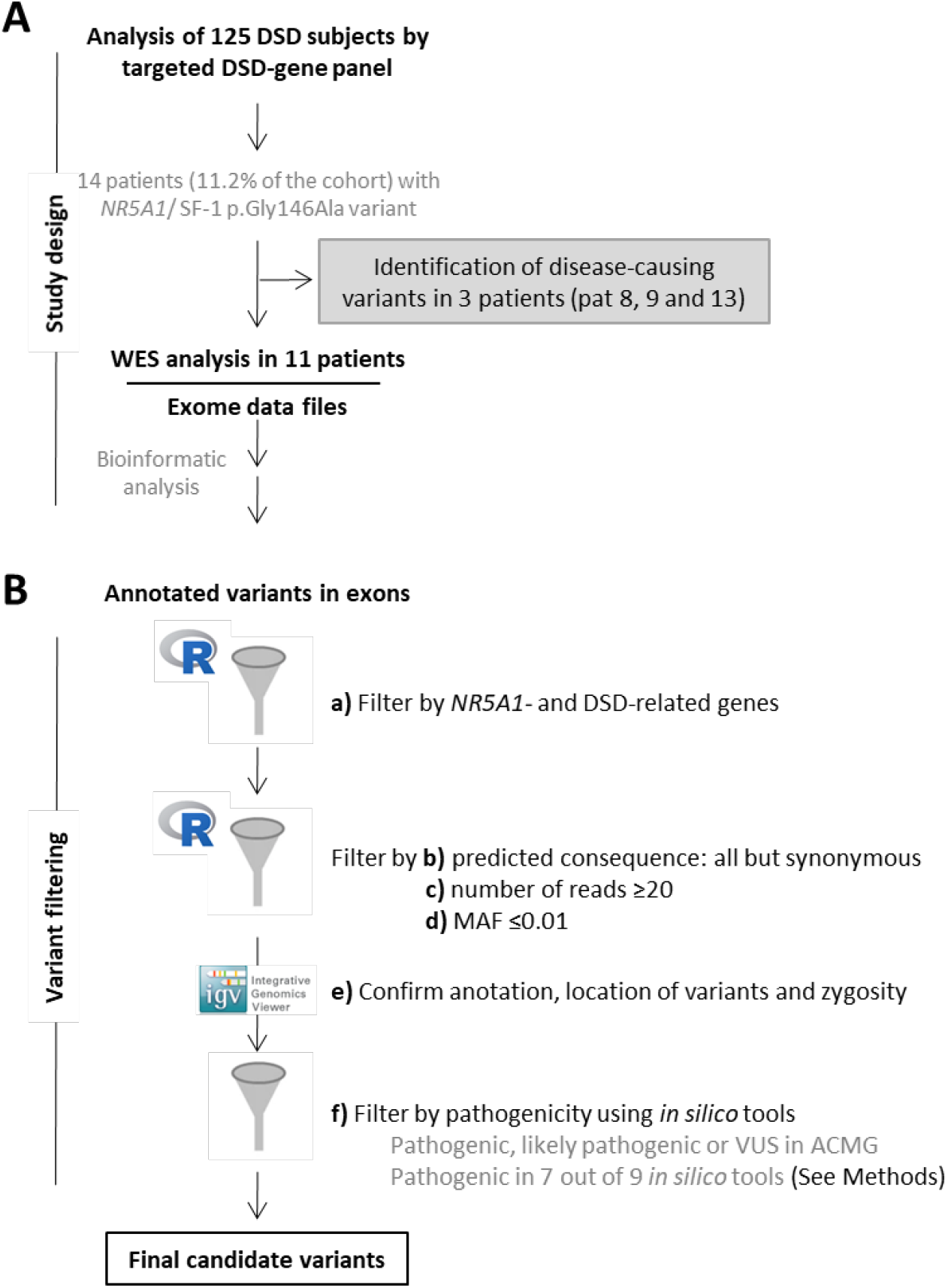
Algorithm of genetic workup. A. Mode of genetic analysis, e.g. panel and whole exome sequencing (WES). Three patients were identified with pathogenic variants in LHCGR, WT1 and AR by panel analysis and were not further analyzed by whole-exome sequencing (WES). B. Filtering algorithm of genetic data. Steps used for variant filtering after WES of eleven DSD patients harboring the NR5A1/SF-1 p.Gly146Ala variant are depicted (letters a to f). Final candidates and their possible impact are listed and characterized in Table 2 and Supplementary Table 3.

These steps included the filtration of WES data by a DSD- and *NR5A1*-related gene list. These disease-tailored lists were updated from previously reported work (DSD-gene list, N=584; *NR5A1*-related gene list, N=628) (22, 32) (Figure 1 Step a). Then, we kept the resulting variants with all type of predicted consequences (e.g. nonsynonymous, frameshift deletion, stop/gain), but synonymous, and with a number of reads above or equal to 20 (33) (Figure 1 steps b and c). Next, we filtered to include variants with a MAF≤0.01 according to gnomAD (v3.1.2) and depending on the origin and karyotype of the patient (Figure 1 step d). In step e, we confirmed the correct annotation, location of variants and zygosity by checking their alignment data in IGV (Integrative Genomics Viewer). Finally, we predicted the possible effect of the identified variant (see below) (Figure 1 step f). Variants were confirmed by visual examination using the IGV (Integrative Genomics Viewer) browser (34, 35).

**Table 2.** Additional gene variants identified in the DSD patients harbouring the *NR5A1*/SF-1 p.Gly146Ala variant.

| Patient | Chromosome position | Gene (Name) | Variant | Type | dbSNP | Zygosity | Previously reported<sup>a</sup> |
| --- | --- | --- | --- | --- | --- | --- | --- |
| 1 | 4:1807384 | *FGFR3* (Fibroblast Growth Factor Receptor 3) | **c.1633_1634del; p.Cys545Hisfs\*17** | frameshift deletion | ND | het |  |
|  | 5:5232601 | *ADAMTS16* (ADAM Metallopeptidase with Thrombospondin Type 1 Motif 16) | **c.1822_1823del; p.His608\*** | stopgain | ND | het |  |
|  | 19:7184641 | *INSR* (Insulin Receptor) | **c.660_661del; p.Pro220Hisfs\*4** | frameshift deletion | ND | het |  |
| 3 | 2:120989442 | *GLI2* (GLI Family Zinc Finger 2) | c.3528G>T; p.Gln1176His | nonsynonymous SNV | rs139686081 | het |  |
|  | 8:60743055 | *CHD7* (Chromodomain Helicase DNA Binding Protein 7) | **c.1623C>A; p.His541Gln** | nonsynonymous SNV | ND | het |  |
|  | 11:77181567 | *MYO7A* (Myosin VIIA) | **c.2882G>A; p.Gly961Asp** | nonsynonymous SNV | rs199575418 | het |  |
|  | 12:47865148 | *VDR* (Vitamin D Receptor) | **c.176C>T; p.Thr59Ile** | nonsynonymous SNV | rs145002466 | het |  |
| 6 | 10:33330774 | *NRP1* (Neuropilin 1) | c.182C>A; p.Pro61Gln | nonsynonymous SNV | ND | het |  |
| 8 | 2:48698724 | *LHCGR* (Luteinizing Hormone/Choriogonadotropin Receptor) | **c.757T>C; p.Ser253Pro** | nonsynonymous SNV | ND | hom |  |
| 9 | 11:32428517 | *WT1* (WT1 Transcription Factor) | c.764T>A; p.Met255Lys | nonsynonymous SNV | rs377573993 | het |  |
| 10 | 9:114275766 | *COL27A1* (Collagen Type XXVII Alpha 1 Chain) | **c.3715C>T; p.Arg1239Trp** | nonsynonymous SNV | rs143724625 | het |  |
|  | 15:41564270 | *TYRO3* (TYRO3 Protein Tyrosine Kinase) | **c.666_667insCACTGCCTGCAGCCC CCTTCAACATCACC; p.Ala223Hisfs\*21** | frameshift insertion | ND | het |  |
| 11 | 16:984721 | *SOX8* (SRY-Box Transcription Factor 8) | **c.676A>C; p.Thr226Pro** | nonsynonymous SNV | ND | het |  |
| 12 | 7:75985941 | *POR* (Cytochrome P450 Oxidoreductase) | **c.1679C>T; p.Thr560Met** | nonsynonymous SNV | rs574694698 | het |  |
|  | 16:2114399 | *PKD1* (Polycystin 1, Transient Receptor Potential Channel Interacting) | **c.2624C>T; p.Pro875Leu** | nonsynonymous SNV | ND | het |  |
|  | 16:30737182 | *SRCAP* (Snf2 Related CREBBP Activator Protein) | **c.7142G>A; p.Arg2381His** | nonsynonymous SNV | rs765139685 | het |  |
|  | 17:72123563 | *SOX9* (SRY-Box Transcription Factor 9) | **c.710dup; p.Pro238Thrfs\*14** | frameshift insertion | ND | het | Campomelic dysplasia (75) |
| 13 | X:67721837 | *AR* (Androgen Receptor) | **c.2323C>T; p.Arg775Cys** | nonsynonymous SNV | rs137852562 | hemi | AIS (66) |
| 14 | 11:77194460 | *MYO7A* (Myosin VIIA) | **c.4259G>A; p.Arg1420His** | nonsynonymous SNV | rs568337942 | het |  |
|  | 16:984739 | *SOX8* (SRY-Box Transcription Factor 8) | **c.694A>C; p.Lys232Gln** | nonsynonymous SNV | rs1596200787 | het |  |

AIS, androgen insensitivity syndrome; Hemi, hemizygous; Het, heterozygous; Hom, homozygous; ND, not determined; POI, primary ovarian insufficiency.
Variants classified as pathogenic, likely pathogenic or as of unknown significance according to the ACMG (American College of Medical Genetics) are highlighted in bold.
<sup>a</sup>Previously associated disease to the specific variant identified in this work.

For the targeted DSD-gene panel analysis, preparation of the libraries and sequencing have been described elsewhere (24). For variant filtration after panel analysis, same steps b to f were followed (Figure 1B).

### *In silico* analyses and variant classification

We predicted the possible effect of identified nonsynonymous genetic variants on the structure and function of the protein using Polyphen-2, (Polymorphism Phenotyping v2), Panther (Protein ANalysis THrough Evolutionary Relationships), SNPs and GO, CADD (Combined Annotation Dependent Depletion) and the calibrated scores given by VarSome (36) for Revel (Rare Exome Variant Ensemble Learner), SIFT (Scale-invariant feature transform), Provean (Protein Variation Effect Analyzer), Mutation taster and M-CAP (Mendelian Clinically Applicable Pathogenicity) (see Supplementary table 3). Variants were classified for pathogenicity according to the standards and guidelines of the ACMG (6) using VarSome. We considered variants as candidates when classified as pathogenic, likely pathogenic or as VUS by the ACMG criteria or when classified as pathogenic or VUS by at least 7 out of 9 prediction programs. Previously reported clinical associations were searched in ClinVar and HGMD databases. In addition, the literature (e.g. PubMed) was searched for evidence of relationship with DSD, sex development and the specific clinical phenotype of each study subject. We explored the possible disease-causing variants’ combinations using ORVAL (Oligogenic Resource for Variant AnaLysis) (37).

## Results and Discussion

In a random cohort of 125 subjects with a DSD, we identified the *NR5A1*/SF-1 p.Gly146Ala variant in 14 individuals (11.2%). The prevalence in 46,XY DSD subjects was 10.1% (10/99), and was in line with previously reported prevalence in this population (15, 16). Prevalence was higher in 46,XX DSD (4/24, 16.7%). Of the 14 studied subjects, five were homozygous and nine heterozygous for the *NR5A1*/SF-1 p.Gly146Ala variant. The phenotype of the individuals ranged from typical for karyotype to mild and severe atypical in 46,XY as well as opposite sex in both 46,XY and 46,XX (Figure 2). Patients were of African (8/14), Spanish (4/14) and Asian (2/14) origin. A summary of the clinical and biochemical characteristics of the patients is given in Table 1 and Supplementary table 2. An overview of the identified genes of our study subjects that likely play a role for the DSD phenotype in a concerted way is given in Figure 3. In this Figure 3 the identified variants are shown within the network of established genes of sex determination and differentiation.

**Figure 2.**
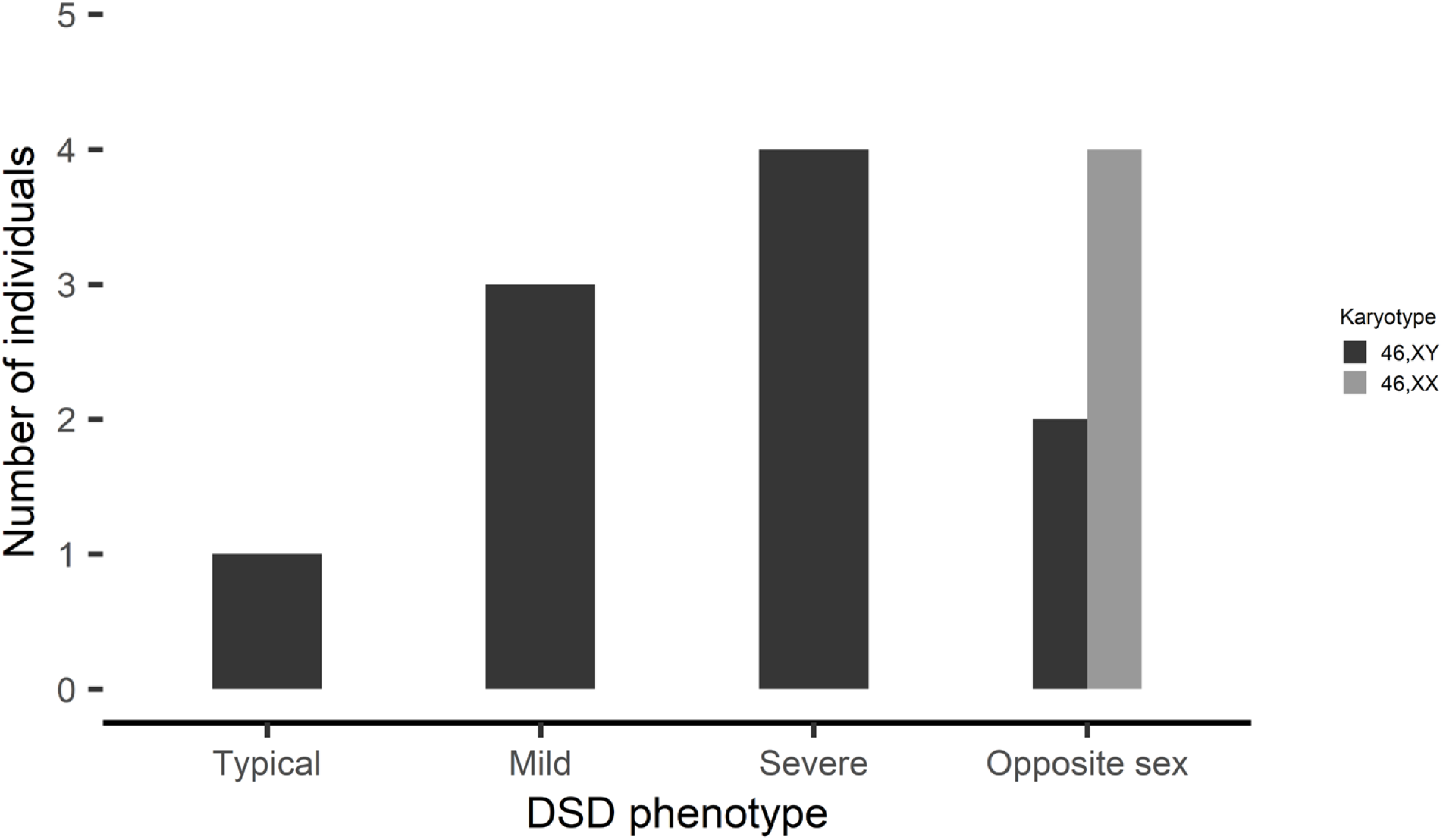
External genital phenotype of the 14 DSD patients harboring the NR5A1/SF-1 p.Gly146Ala variant shown with respect to their karyotype.

**Figure 3.**
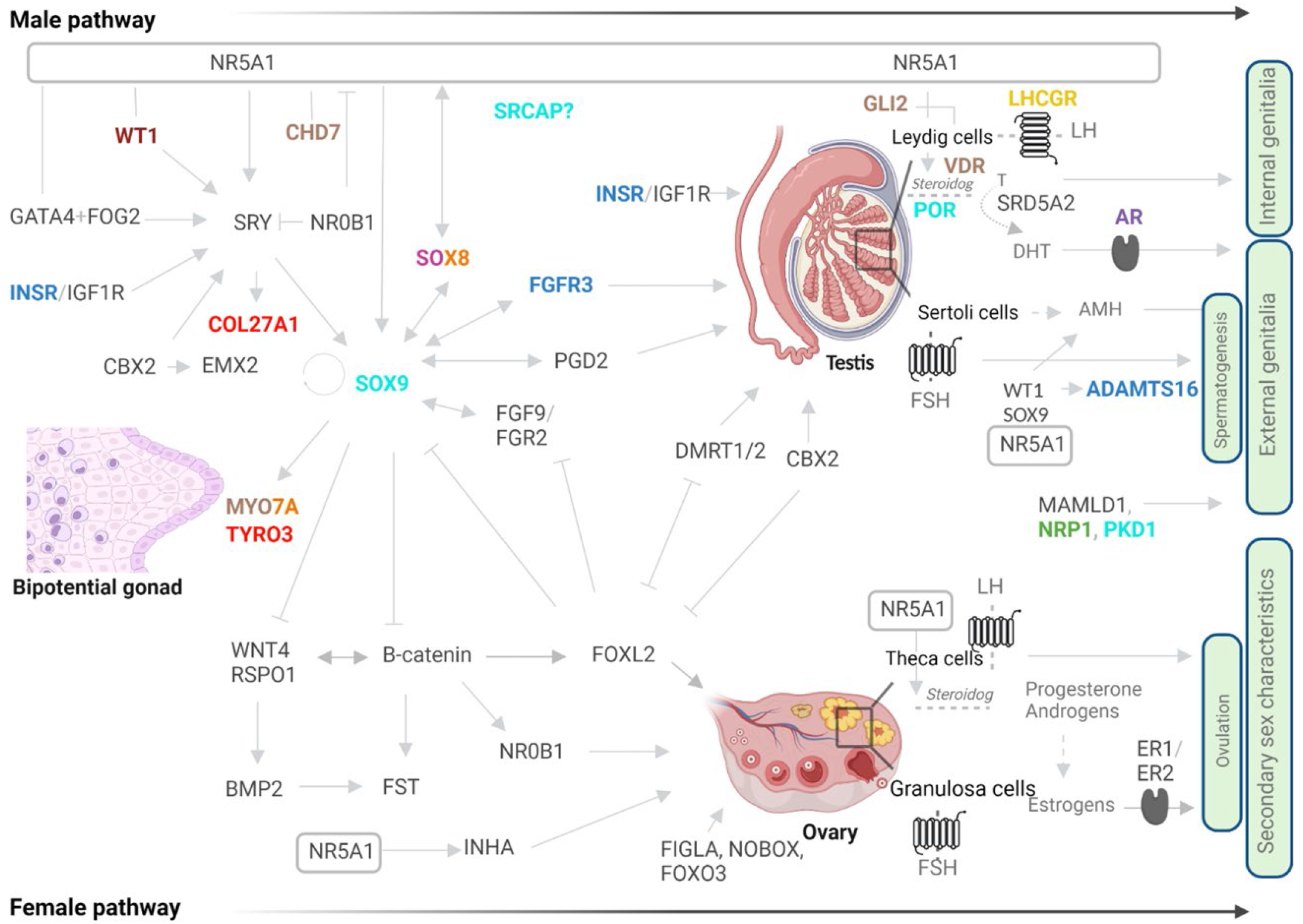
Genetic variants identified in 14 DSD patients harboring the *NR5A1*/SF-1 p.Gly146Ala variant illustrated with respect to the known pathways of male and female sex determination and differentiation. The scheme shows an overview of involved genes and their currently assumed relationship to sexual development. Genes with variants identified by whole exome sequencing in the patients have specific colors. In dark blue: candidate genes in patient 1; in brown: candidate genes in patient 3; in green: candidate genes in patient 6; in yellow: candidate genes in patient 8; in dark red: candidate genes in patient 9; in red: candidate genes in patient 10; in pink: candidate genes in patient 11; in light blue: candidate genes in patient 12; in purple: candidate genes in patient 13; in orange: candidate genes in patient 14; in dark grey: known genes involved in sexual development. Interrogation mark (?): function/timing/location is not clear; arrows: activation; inhibitors: inhibition; lines: interaction/partnership; dashed lines/arrows: hormone production.

NGS performed in DSD individuals harboring the p.Gly146Ala variant in *NR5A1*/SF-1 revealed several deleterious/candidate variants that potentially explain the phenotype of the patients. Overall, we identified either a known pathogenic DSD variant or one to four potentially deleterious/candidate variants in 10 out of the 14 DSD individuals analyzed. A detailed summary of identified variants in other DSD-related genes is shown in Table 2 (further details in Supplementary tables 3 and 4). In three patients we detected variants in known DSD-causing genes with our targeted gene panel, e.g. *LHCGR, WT1*, and *AR*. In 11 patients WES was performed and variants were filtered by candidate gene lists (Figure 1). Overall, the NGS analysis identified 63 variants categorized as (likely) pathogenic or VUS in 57 different genes, however further review of evidence of correlation with DSD, sex development and phenotype of each patient with literature reduced this number to 20 potentially deleterious/candidate variants in 18 genes in 10 subjects. In nine 46,XY DSD individuals 1-4 variants were found in a total of 16 genes, while one 46,XX DSD person revealed two variants in two different genes (Table 2). All variants, identified either by gene panel or WES, but one (e.g. *LHCGR*), were detected in heterozygosis or hemizygosis. Details of the rejected variants are given in Supplementary table 4.

In **patient 1** two frameshift deletions in genes *FGFR3* (c.1633_1634del; p.Cys545Hisfs*17) and *INSR* (c.660_661del; p.Pro220Hisfs*4) and a stop-gain variant in *ADAMTS16* (c.1822_1823del; p.His608*) were found and predicted to be likely pathogenic by the ACMG criteria. *FGFR3* is essential for testicular development in humans (38), while *INSR* and *ADAMTS16* are needed in murine genitourinary development and testicular differentiation, respectively (39, 40). Variants in *ADAMTS16* have also been reported in heterozygosis in two 46,XY females with complete gonadal dysgenesis and a 46,XY DSD patient with ambiguous genitalia (41). Testing for a digenic combination network with ORVAL showed no variant interaction between *ADAMTS16* and *FGFR3*.

We detected four heterozygous missense variants in four genes in **patient 3**. These were *GLI2* (c.3528G>T; p.Gln1176His), *CDH7* (c.1623C>A; p.His541Gln), *MYO7A* (c.2882G>A; p.Gly961Asp) and *VDR* (c.176C>T; p.Thr59Ile). The variant in *GLI2* (c.3528G>T; p.Gln1176His) was rated as pathogenic by most of the *in silico* prediction tools and variants in additional genes were rated as VUS when analyzing according to pathogenicity guidelines. Variants in *GLI2* have been described in syndromic DSD patients together with short stature, primary hypogonadism, heart and craniofacial anomalies and 46,XY gonadal dysgenesis (42), as well as in 46,XY non-syndromic DSD manifesting with female external genitalia or with ambiguous genitalia (23, 43, 44). Variants in *CHD7* have been previously associated with a broad range of 46,XY DSD phenotypes, including males with hypospadias or micropenis and individuals with female external genitalia (27, 45). *MYO7A* is overexpressed in male supporting cells during gonadal development (46) and has been shown to be a SRY and SOX9 target gene (47), but, in DSD individuals it has been identified only in combination with *MAMLD1* (44, 48). Finally, *VDR* plays a dominant role in male fertility as Vdr^-/-^ mice show abnormal gonads in both sexes and variable reproductive phenotypes such as reduced sperm count (49). In humans, two polymorphisms in *VDR* were associated with female idiopathic infertility only (50). Fertility of patient 3 has not been assessed yet, and we cannot exclude a role of the *VDR* variant in his DSD phenotype. Network analysis by ORVAL predicts a pathogenic gene network between *CHD7, MYO7A* and *GLI2* (Supplementary figure 1A). A heterozygous missense c.182C>A; p.Pro61Gln variant in Neuropilin 1 (*NRP1)* gene was found in **patient 6**. NRP1 interacts with Sema3A which is essential for the development of the GnRH neuron system (51). Loss of Sema3a (Semaphorin 3A) signaling in mice results in reduced gonadal size and recapitulates the features of Kallmann syndrome (52). In humans, variants in *NRP1* have been identified in a 46,XY DSD subject with female external genitalia (45) and a 46,XY male presenting with penoscrotal hypospadias, in whom other genetic variants were identified, among them a variant in *MAMLD1*, a known DSD-related gene (48).

In 46,XY **patient 8** with a phenotype of opposite sex a homozygous, inactivating variant in *LHCGR* (c.757T>C; p.Ser253Pro) was found. This variant has been previously reported to severely reduce the signal transduction activity of the LH receptor and therefore leads to the complete form of Leydig cell hypoplasia (LCH) as seen in patient 8 (53).

A novel, likely pathogenic p.Met255Lys (c.764T>A) missense variant in exon 2 of *WT1* was found in **patient 9**, who had precocious puberty and presented with a pituitary adenoma and hearing loss. *WT1* is associated with 46,XY gonadal dysgenesis in syndromes including Denys-Drash or Frasier (54), but more recently *WT1* variants have also been detected in individuals with isolated DSD, such as ambiguous genitalia (45, 55, 56) or gonadal dysgenesis. WT1 acts as a transcriptional repressor or activator on many target genes, such as *LHB* (Luteinizing Hormone Subunit Beta), which encodes the beta subunit of LH (luteinizing hormone), responsible for testosterone production in males during puberty. Variants of *WT1* may therefore affect the transcription of LHβ and thereby lead to altered puberty timing as observed in patient 9. In fact, in mouse cell models, WT1 regulates LHB transcription and two splice variants (-KTS/+KTS) were already shown to have positive and negative roles in this regulation (57).

A missense variant in *COL27A1* (c.3715C>T; p.Arg1239Trp) and a frameshift insertion in *TYRO3* (c.666_667insCACTGCCTGCAGCCCCCTTCAACATCACC; p.Ala223HisfsTer21) were found in **patient 10**. Both variants were categorized as VUS and were detected in heterozygosis. In mice, *Col27a1* is highly expressed in XY somatic supporter cells compared to XX during the earliest stages of gonad development (58). *Col27a1* has been identified as a SRY target gene in the embryonic mouse gonads at E11.5 by ChIP-Chip experiments (47). Similarly, *Tyro3* is overexpressed in male somatic cells (59), and is regulated by SOX9 (47). Protein truncating variants of *TYRO3* were found in individuals with idiopathic hypogonadotropic hypogonadism establishing a role of this gene in reproductive development (60). Taken together, the data suggest that both *COL27A1* and *TYRO3* genes are part of the genetic network underlying the early stages of mammalian fetal gonadal development, and thus genetic changes are likely causing the ovotesticular DSD phenotype in patient 10. However, a gene interaction between *COL27A1* and *TYRO3* was not predicted by ORVAL.

In 46,XY **patient 11** with a distal hypospadias, one missense variant in the *SOX8* (c.676A>C; p.Thr226Pro) gene was detected. It was identified in heterozygosis and was classified as VUS. *SOX8* is involved in early testis determination (61). *SOX8* gene variants are associated with a range of phenotypes including 46,XY DSD and human reproductive anomalies in males and females (62). Single-nucleotide variants (SNV) associated with 46,XY gonadal dysgenesis are mostly located in the HMG-box of SOX8 (43), while those reported in infertile patients flank the HMG-box or localize to one of the transactivation domains (TA) (62). However, more recently, a missense variant in the TA2 region of SOX8 was identified in a 46,XY patient with gonadal dysgenesis (43). The novel c.676A>C; p.Thr226Pro variant is located in the first TA of the protein. *In vitro* studies have shown impaired cellular localization in some mutant proteins located in this functional domain of SOX8. Therefore, this missense variant likely explains the genital phenotype observed in patient 11. Biochemical assessment (age range 11-15) of the HPG axis was normal and pubertal development was ongoing (Tanner 3-4).

Four heterozygous VUS or likely pathogenic variants were identified in **patient 12** with a severe 46,XY DSD phenotype. These were *POR* (c.1679C>T; p.Thr560Met), *PKD1* (c.2624C>T; p.Pro875Leu), *SRCAP* (c.7142G>A; p.Arg2381His) and *SOX9* (c.710dup; p.Pro238Thrfs*14). The involvement of *POR* and *SOX9* in sexual development is well known and several sequence variants have been described in 46,XY DSD patients (27, 47, 63). *Pkd1* is critical for epididymal epithelium development and for maintaining mice male reproductive tract (64). *PKD1* variants have not been related to DSD yet, but they cause autosomal dominant polycystic kidney disease (ADPKD), which involves reproductive tract abnormalities and infertility in males (65). Therefore, a role of *PKD1* variants in DSD seems possible. Likewise, the role of *SRCAP* in sex differentiation and development is unknown. However, this is the second 46,XY DSD patient in whom a gene variant is identified (42). According to ORVAL analysis, oligogenic pathogenicity is predicted by combination of variants in a gene network including *POR, PKD1* and *SRCAP* (Supplementary figure 1B).

In **patient 13**, we identified an *AR* variant (c.2323C>T; p.Arg775Cys) previously reported in a patient with Complete Androgen Insensitivity Syndrome (CAIS) (66). Because the patient presented with a typical CAIS phenotype, it seems plausible that this hemizygous *AR* variant is fully responsible for the DSD.

**Patient 14**, with a severe 46,XY phenotype, harbored two heterozygous missense variants in *MYO7A* and *SOX8* genes. Both were categorized as VUS by the ACMG. As in patient 11, the *SOX8* variant (c.694A>C; p.Lys232Gln) was also located in the TA1 domain of the protein. However, the phenotype of patient 14 was more severe, either caused by the *SOX8* variant alone or due to the digenic effect together with *MYO7A*. Importantly, the combination of variants in *SOX8* and *MYO7A* is predicted as disease-causing by ORVAL (Supplementary figure 1C). The combination of variants in *MYO7A* and *SOX8* in DSD was reported previously (44, 48), and suggests that the broad phenotype observed in DSD individuals might be explained by oligogenic origin (67).

In four patients carrying the heterozygous p.Gly146Ala *NR5A1* variant, the WES and specific data analysis revealed no other candidate genes explaining their DSD phenotype. Of these patients 2, 4 and 5 had a 46,XX karyotype and an opposite genital phenotype, and were assigned as males at birth, whereas patient 7 presented with a severe 46,XY DSD. None had other organ anomalies. Although NGS has facilitated the identification of the underlying genetic defects of DSD, about 50% of individuals with a 46,XY DSD remain without a molecular diagnosis with currently used methods (27). We used WES in our study, while other genetic studies also search for variants in noncoding, regulatory or intronic regions by whole genome sequencing (WGS). But even when using WGS, a considerable number of patients are reportedly unsolved (68). Thus, other factors such as environmental factors or epigenetic regulators have been suggested playing a role (68, 69). In addition, oligogenic or even polygenic inheritance might be considered for explaining the broad phenotypes seen in some individuals with a DSD (23-29, 48, 67, 70-73). In early days of genetic workup of DSD, patients were studied by candidate Sanger sequencing. In 46,XY DSD subjects typical candidates were the *AR, SRD5A2* and *NR5A1*/SF-1; and once a genetic variant was found, additional genes were not tested. Thus, some DSD patients that have been tested by the candidate approach may not have a correct diagnosis and need to be retested by NGS.

In conclusion, NGS genetic analysis of DSD individuals carrying the p.Gly146Ala variant of the *NR5A1*/SF-1 gene revealed variants in other genes (likely) explaining their phenotype. These gene variants were either found in established DSD genes, were previously described or novel, and were (likely) disease-causing either in a monogenic or in a suggested oligogenic fashion. Although we were not able to find causal genetic variants in four out of 14 DSD individuals carrying the *NR5A1*/SF-1 p.Gly146Ala, our study supports the claim that this *NR5A1*/SF-1 variant is a benign polymorphism that does not play a role in the pathogenesis of DSD. Therefore, we strongly recommend reanalyzing DSD patients of whom phenotype has been thought to be explained by this variant in order to find the real underlying genetic cause.

## Supporting information

S1 Table. Genes included in the customized DSD panel and their suggested role in DSD.

## Data Availability

All data produced in the present study are available upon reasonable request to the authors

## Acknowledgement

The authors thank the patients and their families for participating in our research.

## Competing interests

The authors have declared that no competing interests exist.

## Funding

This work has been supported by the University of the Basque Country (UPV-EHU) (Spain) (IT1739-22) and by the Swiss National Science Foundation (320030-197725). IM is supported by a Postdoctoral Fellowship Grant from the Education Department of Basque Government (Spain). JA is supported by ASONMEC (Asociacion de Oncologia Medica del Hospital de Cruces) (Spain).

## Supporting information

S1 Table. Genes included in the customized DSD panel and their suggested role in DSD.

S2 Table. Complete description of the phenotype and biochemical data of the DSD patients harbouring the NR5A1/SF-1 p.Gly146Ala variant.

S3 Table 3. Gene variant characterization: allele frequency and disease prediction by ACMG classification and by different in silico programs aSpecific allele frequency for the origin and karyotype of the patient. bCADD phred score >20 indicates that the variant is predicted to be the 1% most deleterious substitution that you can do to the human genome.

S4 Table. List of rejected variants identified in the DSD patients harbouring the NR5A1/SF-1 p.Gly146Ala variant. Variants were discarded after filtering due to weak relation to DSD, zygosity or absence of correspondence to the phenotype.

S1 Figure. Potential oligogenic interaction networks of DSD- and NR5A1-related genes identified in specific DSD individuals harbouring the NR5A1/SF-1 p.Gly146Ala variant. Networks were identified for patients 3, 12 and 14 respectively. To search for potential oligogenic disease networks, the Oligogenic Resource for Variant AnaLysis (ORVAL, https://orval.ibsquare.be/) was used. Nodes represent genes and edges connect two genes only, if between them there is at least one candidate disease-causing variant combination predicted by VarCoPP. The colour of the edge represents the pathogenicity score for that pair of genes. This score is represented in a colour range from brown (higher pathogenicity score) to yellow (lower pathogenicity score).

## Notes

### Competing Interest Statement

The authors have declared no competing interest.

### Author Declarations

The study was approved by the ethics committee for clinical research of Euskadi (CEIC-E), Spain

