## Supplementary material for "The *NR5A1/SF-1* variant p.Gly146Ala cannot explain the phenotype of individuals with a difference of sex development": S1 Table. Genes included in the customized DSD panel and their suggested role in DSD.

Supplemental Material

Supplementary table 1. Genes included in the customized DSD panel and their suggested role in DSD.

| **Gene (*locus*)** | **Alias** | **Transcript** | **Role in DSD** |
|---|---|---|---|
| *AMH* (19p13.3) | Anti-Mullerian Hormone | NM_000479.3 | G diff |
| *AMHR2* (12q13.13) | Anti-Mullerian Hormone Receptor Type 2 | NM_020547.2 | G diff |
| *AR* (Xq12) | Androgen Receptor | NM_000044.3 | G diff |
| *ATRX* (Xq21.1) | ATP-Dependent Helicase ATRX | NM_000489.4 | G diff |
| *BMP15* (Xp11.22) | Bone Morphogenetic Protein 15 | NM_005448.2 | G det |
| *CBX2* (17q25.3) | Chromobox homolog 2 | NM_005189.2 | G det |
| *CYP11A1* (15q24.1) | Cytochrome P450 Family 11 Subfamily A Member 1 | NM_000781.2 | G diff |
| *CYP11B1* (8q24.3) | Cytochrome P450 Family 11 Subfamily B Member 1 | NM_000497.3 | G diff |
| *CYP17A1* (10q24.32) | Cytochrome P450 Family 17 Subfamily A Member 1 | NM_000102.3 | G diff |
| *CYP19A1* (15q21.2) | Cytochrome P450 Family 19 Subfamily A Member 1 | NM_000103.3 | G diff |
| *CYP21A2* (6p21.33) | Cytochrome P450 Family 21 Subfamily A Member 2 | NM_000500.7 | G diff |
| *DHH* (12q13.12) | Desert Hedgehog | NM_021044.2 | G det |
| *DMRT1* (9p24.3) | Doublesex And Mab-3 Related Transcription Factor 1 | NM_021951.2 | G det |
| *DMRT2* (9p24.3) | Doublesex And Mab-3 Related Transcription Factor 2 | NM_181872.4 | G Dev |
| *ESR1* (6q25.1-q25.2) | Oestrogen Receptor 1, Nuclear Receptor Subfamily 3 Group A Member 1 | NM_001122740.1 | G diff |
| *ESR2* (14q23.2-q23.3) | Oestrogen Receptor 2, Nuclear Receptor Subfamily 3 Group A Member 2 | NM_001437.2 | G diff |
| *FGF9* (13q12.11) | Fibroblast Growth Factor 9 | NM_002010.2 | G det |
| *FOXL2* (3q22.3) | Forkhead Box L2 | NM_023067.3 | G det |
| *FOXO3* (6q21) | Forkhead Box O3 | NM_001455.3 | G det |
| *FSHR* (2p16.3) | Follicle Stimulating Hormone Receptor | NM_000145.3 | CHH |
| *GATA4* (8p23.1) | GATA Binding Protein 4 | NM_002052.3 | G det |
| *HARS2 (5q31.3)* | Histidyl-TRNA Synthetase 2, Mitochondrial | NM_012208.3 | G diff |
| *HSD17B3 (*9q22.32) | Hydroxysteroid 17-Beta Dehydrogenase 3 | NM_000197.1 | G diff |
| *HSD17B4* (5q23.1) | Hydroxysteroid 17-Beta Dehydrogenase 4 | NM_000414.3 | G diff |
| *HSD3B2* (1p12) | Hydroxy-Delta-5-Steroid Dehydrogenase, 3 Beta and Steroid Delta-Isomerase 2 | NM_000198.3 | G diff |
| *INHA* (2q35) | Inhibin Subunit Alpha | NM_002191.3 | G det |
| *INSL3* (19p13.11) | Insulin Like 3 | NM_001265587.1 | G diff |
| *KISS1* (1q32.1) | Kisspeptin-1 | NM_002256.3 | CHH |
| *KISS1R* (19p13.3) | KISS1 Receptor | NM_032551.4 | CHH |
| *LHCGR* (2p16.3) | Luteinizing Hormone/Choriogonadotropin Receptor | NM_000233.3 | G diff |
| *MAMLD1* (Xq28) | Mastermind Like Domain Containing 1, CXorf6 | NM_001177465.2 | G det |
| *MAP3K1* (5q11.2) | Mitogen-Activated Protein Kinase Kinase Kinase 1 | NM_005921.1 | G det |
| *NR0B1* (Xp21.2) | Nuclear Receptor Subfamily 0 Group B Member 1 | NM_000475.4 | G det |
| *NR5A1* (9q33.3) | Nuclear Receptor Subfamily 5 Group A Member 1 | NM_004959.4 | G det |
| *POR* (7q11.23) | Cytochrome P450 Oxidoreductase | NM_000941.2 | G diff |
| *PSMC3IP* (17q21.2) | Proteasome 26S ATPase Subunit 3-Interacting Protein | NM_016556.3 | G det |
| *RSPO1* (1p34.3) | R-Spondin 1 | NM_001242908.1 | G det |
| *RXFP2* (13q13.1) | Relaxin Family Peptide Receptor 2, GREAT, LGR8 | NM_130806.3 | G diff |
| *SOX3* (Xq27.1) | SRY (Sex Determining Region Y)-Box 3 | NM_005634.2 | G det |
| *SOX9* (17q24.3) | SRY (Sex Determining Region Y)-Box 9 | NM_000346.3 | G det |
| *SRD5A2* (2p23.1) | Steroid 5 Alpha-Reductase 2 | NM_000348.3 | G diff |
| *SRY* (Yp11.2) | Sex Determining Region Y | NM_003140.2 | G det |
| *STAR* (8p11.23) | Steroidogenic Acute Regulator | NM_000349.2 | G diff |
| *TSPYL1* (6q22.1) | Testis-Specific Y-Encoded-Like Protein 1 | NM_003309.3 | G det |
| *WNT4* (1p36.12) | Wingless-Type MMTV Integration Site Family, Member 4 | NM_030761.4 | G det |
| *WT1* (11p13) | Wilms Tumour 1 | NM_024426.4 | G det |
| *WWOX* (16q23.1-q23.2) | WW Domain Containing Oxidoreductase | NM_016373.2 | G det |
| *ZFPM2* (8q23.1) | Zinc Finger Protein, FOG Family Member 2, Friend Of GATA 2 | NM_012082.3 | G det |

CHH, central causes of hypogonadism; G det, gonadal determination; G diff, gonadal differentiation.

Supplementary table 2. Complete description of the phenotype and biochemical data of the DSD patients harbouring the *NR5A1*/SF-1 p.Gly146Ala variant.

| Patient | Phenotype | Age range at evaluation | Adrenal steroidogenesis | | | | | Gonadal function | | | | | |
|---|---|---|---|---|---|---|---|---|---|---|---|---|---|
| | | | ACTH (pg/mL) | Cortisol (μg/dL) | 17OHP4 (ng/mL) | DHEA-S (ng/mL) | Δ4-A (ng/mL) | Testosterone (ng/dL) | FSH (U/L) | LH (U/L) | E2 (pg/mL) | DHT (ng/ml) | AMH (ng/mL) |
| 1 | 11-15y, after treatment, mature penis and scrotum, pubarche IV | 0-5y | <25 | | | | | **34.7** | | | | | |
| 2 | 6-10y, micropenis, scrotal hypospadias, bifid scrotum. | 6-10y | | | **<0.1** | <180 | | <10/44.1* | | | | **0.3/**0.1* | |
| 3 | 6-10y, micropenis (2cm), scrotal hypospadias. | 6-10y | | | | | | 80* | | | | N* | |
| 4 | 6-10y, curved penis (2cm), scrotal hypospadias, bifid scrotum, right inguinal hernia. | 6-10y | | | 0.2 | <180 | <0.3 | <10/125* | <1.5 | <1.5 | | <0.1/0.1* | |
| 5 | 0-5y, rudimentary penis (<0.5cm), pubarche I. | 0-5y | | | | | **0.5/<0.3*** | **14/59*** | 1,3 | <0.1 | | **0.1/0.1*** | 8.8 |
| 6 | 0-5y, curved penis, scrotal hypospadias. | 0-5y | | 17.7 | | <150 | <0.3/<0.3* | <10/110.4* | | | | **<0.1/<0.1*** | |
| 7 | 6-10y, curved penis (4.4-4.5cm), scrotal hypospadias, cryptorchidism and bifid scrotum | 6-10y | | 11 | 0.4 | 270 | <0.3/<0.3* | <10/263.4* | 0.8 | <0.1 | | 0.1/0.3* | 22.8 |
| 8 | 6-10y, right inguinal hernia. At puberty, primary amenorrhea, ovarian and uterine agenesis. 21-25y, left inguinal hernia. 31-35y, female external genitalia, thelarche II-III, erectile organ is buried, one opening for urethra and vagina. | 31-35y | 27.9 | 21.3 | 0.2 | 1390 | 1.3 | **19** | **67.0** | **48.5** | 5 | | |
| 9 | 6-10y, penis 8cm, testes 10mL, pubarche II. | 6-10y | | | 1.4 | 0.6 | | **434.7** | 4.6 | 4.6 | 22 | | |

| | | | | | | | | | | | | |
|---|---|---|---|---|---|---|---|---|---|---|---|---|
| 10 | 0-5y, curved penis, scrotal hypospadias, atrophic scrotum, non-palpable testes. | 6-10y | | 0.3 | 1100 | | <20 | 0.5 | <10 | | | |
| 11 | 0-5y, distal hypospadias, surgery. 11-15y, after treatment, penis (5.2cm) buried in fat, testes 10-12ml, pubarche III. | 0-5y | 13.9 | 0.1 | <150 | <0.3/<0.3* | **<10/**72.8* | **3** | <0.1 | | **<0.1** | 7 |
| 12 | 0-5y, left scrotal hypoplasia. Orchidopexy. 11-15y, penis 3.5-4cm, testes 2-4ml, gynecomastia, pubarche I. | 6-10y | | | | <0.3 | <10 | 1.3 | <0.1 | | 0.1 | 193 |
| 13 | 11-15y, primary amenorrhea. 41-45y, absence of developed breast, pubic and axillary hair. | ND | | | | | | | | | | |
| 14 | 0-5y, micropenis, fused labia minora, non-palpable gonads, tight vagina. | 0-5y | 21.4 | **8.2** | 247 | **3.2** | 1.5 | **17.3** | **17.1** | 16.2 | | |

ACTH, adrenocorticotropic hormone; AMH, anti-Müllerian hormone; DHEA-S, dehydroepiandrosterone sulfate; DHT, dihydrotestosterone; E2, estradiol; FSH, follicle-stimulating hormone; LH, luteinizing hormone; N, normal; ND, not determined; PRL, prolactin; P4, progesterone; Y, years; Δ4-A, delta 4-androstenedione; 17OHP4, 17-hydroxy-progesterone. (*) Values after stimulation with hCG or ACTH. Out of range values for karyotypic sex and age are given in bold.

Supplementary table 3. Gene variant characterization: allele frequency and disease prediction by ACMG classification and by different *in silico* programs.

| Patient | Gene | Variant | Exon | GnomAD (Overall/Specific[a]) | ClinVar | ACMG classification (Criteria) | SIFT | Provean | Polyphen | Mutation Taster | Panther | SNPs and Go | M-CAP | CADD[b] | REVEL |
|---|---|---|---|---|---|---|---|---|---|---|---|---|---|---|---|
| 1 | *FGFR3* | p.Cys545Hisfs*17 | 12 | ND/ND | ND | LP (PVS1,PM2) | ND | ND | ND | ND | ND | ND | ND | ND | ND |
| | *ADAMTS16* | p.His608* | 12 | ND/ND | ND | LP (PVS1,PM2) | ND | ND | ND | ND | ND | ND | ND | ND | ND |
| | *INSR* | p.Pro220Hisfs*4 | 3 | ND/ND | ND | LP (PVS1,PM2) | ND | ND | ND | ND | ND | ND | ND | ND | ND |
| 3 | *GLI2* | p.Gln1176His | 13 | 0.001603/0.004818 | B | B (BP6,BS1,BS2,BP4,BP1) | Path | VUS | Prdam | VUS | Prdam | Dis | B | 22.5 | B |
| | *CHD7* | p.His541Gln | 2 | 0.0001314/0.0001554 | ND | VUS (PM2,PP3) | B | B | B | VUS | ND | Dis | VUS | ND | B |
| | *MYO7A* | p.Gly961Asp | 23 | 0.0005718/0.002384 | VUS | VUS (PM2) | B | Path | Psdam | VUS | ND | Dis | VUS | 26.4 | VUS |
| | *VDR* | p.Thr59Ile | 4 | 0.0006569/0.002487 | LB | VUS (PM1,PM2,PP3,BP6) | B | VUS | Psdam | VUS | Prdam | Dis | VUS | 24.8 | VUS |
| 6 | *NRP1* | p.Pro61Gln | 2 | ND/ND | ND | LB (BP1,BP4,PM2) | Path | VUS | Prdam | VUS | Psdam | Dis | B | ND | VUS |
| 8 | *LHCGR* | p.Ser253Pro | 9 | ND/ND | ND | VUS (PM2) | VUS | VUS | Prdam | VUS | Psdam | ND | VUS | ND | VUS |
| 9 | *WT1* | p.Met255Lys | 2 | 0.00004600/0.000 | LB | B (BS1,BS2,BP6,BP4) | Path | VUS | Psdam | B | Prdam | ND | VUS | 25.1 | VUS |
| 10 | *COL27A1* | c.3645_3651+5del | 37 | 0.001025/0.002889 | LB | VUS (PP3,PM2,BP6) | VUS | Path | Prdam | B | Prdam | N | B | 33 | B |
| | *TYRO3* | p.Ala223HisfsTer21 | 5 | ND/ND | ND | VUS (PM2) | ND | ND | ND | ND | ND | ND | ND | ND | ND |
| 11 | *SOX8* | p.Thr226Pro | 3 | 0.0003858/0.0007298 | ND | VUS (PP3,PM2,BP1) | B | Path | Prdam | VUS | Prdam | Dis | Path | ND | Path |
| 12 | *POR* | p.Thr560Met | 14 | 0.00005255/0.0006859 | ND | VUS (PM2) | B | VUS | B | VUS | Prdam | N | Path | 24.4 | Path |
| | *PKD1* | p.Pro875Leu | 11 | ND/ND | ND | VUS (PM2,PP3) | Path | Path | Prdam | VUS | ND | Dis | Path | ND | VUS |
| | *SRCAP* | p.Arg2381His | 34 | 0.000006576/0.000 | ND | VUS (PM2,BP1) | Path | B | Prdam | B | Prben | N | VUS | 26.8 | B |
| | *SOX9* | p.Pro238Thrfs*14 | 3 | ND/ND | VUS | LP (PVS1) | ND | ND | ND | ND | ND | ND | ND | ND | ND |
| 13 | *AR* | p.Arg775Cys | 6 | ND/ND | P | P (PP5,PS3,PP3,PM1) | Path | Path | ND | VUS | Prdam | ND | Path | 28.2 | Path |
| 14 | *MYO7A* | p.Arg1420His | 32 | 0.00001971/0.000 | VUS | VUS (PM2) | B | B | B | VUS | ND | Dis | VUS | 25.6 | VUS |
| | *SOX8* | p.Lys232Gln | 3 | 0.0002685/0.0002686 | ND | VUS (PP3,PM2,BP1) | VUS | VUS | Prdam | VUS | Prdam | Dis | Path | 27.5 | Path |

B, benign; Dam, damaging; DC, disease causing; Dis, disease; LB, likely benign; LP, likely pathogenic; N, neutral; ND, not determined; P, polymorphism; Path, pathogenic; Psdam, possibly damaging; Prben, probably benign; Prdam, probably damaging; VUS, variant of unknown significance.

[a]Specific allele frequency for the origin and karyotype of the patient. [b]CADD phred score >20 indicates that the variant is predicted to be the 1% most deleterious substitution that you can do to the human genome.

For each gene, sequence information is based on: ADAMTS16 (NM_139056.4), AR (NM_000044.3), CHD7 (NM_017780.4), COL27A1 (NM_032888.4), FGFR3 (NM_000142.5), GLI2 (NM_001374353.1), INSR (NM_000208.4), LHCGR (NM_000233.4), MYO7A (NM_000260.4), NRP1 (NM_003873.7), PKD1 (NM_001009944.3), POR (NM_001395413.1), SOX8 (NM_014587.5), SOX9 (NM_000346.4), SRCAP (NM_006662.3), TYRO3 (NM_006293.4), VDR (NM_000376.3), WT1 (NM_024426.4).

Supplementary table 4. List of rejected variants identified in the DSD patients harbouring the *NR5A1*/SF-1 p.Gly146Ala variant. Variants were discarded after filtering due to weak relation to DSD, zygosity or absence of correspondence to the phenotype.

| Patient | Chromosome position | Gene (Name) | Variant | dbSNP | Zygosity | ACMG classification | Previously reported |
|---|---|---|---|---|---|---|---|
| 1 | 3:52834677 | *ITIH3* (Inter-Alpha-Trypsin Inhibitor Heavy Chain 3) | c.1199_1200del;p.Val400Glyfs*2 | ND | het | LP | No |
| | 5:7886651 | *MTRR* (5-Methyltetrahydrofolate-Homocysteine Methyltransferase Reductase) | c.1094C>A;p.Ser365Tyr | rs1293804430 | het | VUS | No |
| | 7:44112925 | *POLM* (DNA Polymerase Mu) | c.1450dup;p.Leu484Profs*2 | ND | het | VUS | No |
| | 7:44801187 | *ZMIZ2* (Zinc Finger MIZ-Type Containing 2) | c.1381dup;p.Met461Asnfs*4 | ND | het | LP | No |
| | 8:89982837 | *NBN* (Nibrin) | c.56T>G;p.Leu19Trp | rs749263651 | het | VUS | No |
| | 9:133732566 | *SARDH* (Sarcosine Dehydrogenase) | c.367G>T;p.Val123Leu | rs1391072810 | het | VUS | No |
| | 15:41191433 | *VPS18* (VPS18 Core Subunit Of CORVET And HOPS Complexes) | c.417_418insA;p.Gln140Thrfs*32 | ND | hom | VUS | No |
| | 16:50334767 | *ADCY7* (Adenylate Cyclase 7) | c.1218_1219del;p.Glu406Aspfs*98 | ND | het | LP | No |
| | 17:31169974 | *NF1* (Neurofibromin 1) | c.563C>A;p.Ala188Glu | ND | het | VUS | No |
| | 22:19968739 | *ARVCF* (ARVCF Delta Catenin Family Member) | c.889_890insG;p.His297Argfs*11 | ND | hom | VUS | No |
| 2 | 1:88962138 | *KYAT3* (Kynurenine Aminotransferase 3) | c.461T>C;p.Leu154Pro | rs75696718 | het | B | No |
| | 3:142553920 | *ATR* (ATR Serine/Threonine Kinase) | c.2437A>G;p.Met813Val | rs769648140 | het | VUS | No |
| | 5:55960477 | *IL6ST* (Interleukin 6 Cytokine Family Signal Transducer) | c.898C>T;p.Arg300Cys | rs141500365 | het | LB | No |
| | 16:68819395 | *CDH1* (Cadherin 1) | c.1681T>C;p.Tyr561His | ND | het | VUS | No |
| | 20:50192055 | *CEBPB* (CCAAT Enhancer Binding Protein Beta) | c.1022C>T;p.Ser341Phe | ND | het | LB | No |
| 4 | 1:88962138 | *KYAT3* (Kynurenine Aminotransferase 3) | c.461T>C;p.Leu154Pro | rs75696718 | het | B | |
| | 6:117394711 | *ROS1* (ROS Proto-Oncogene 1, Receptor Tyrosine Kinase) | c.911del;p.Leu304Tyrfs*7 | rs763595603 | het | LP | No |
| | X:154532945 | *G6PD* (Glucose-6-Phosphate Dehydrogenase) | c.1048G>C;p.Asp350His | rs34193178 | het | LB | G6PDH deficiency (1) |
| 6 | 3:142553920 | *ATR* (ATR Serine/Threonine Kinase) | c.2437A>G;p.Met813Val | rs769648140 | het | VUS | No |
| | 17:74842561 | *GRIN2C* ( Glutamate Ionotropic Receptor NMDA Type Subunit 2C) | c.3575_3576del;p.Leu1192Argfs*86 | ND | het | VUS | No |
| 7 | 1:241857358 | *EXO1* (Exonuclease 1) | c.419_420del;p.Gln140Argfs*10 | rs1491146265 | het | LP | No |
| | 6:108663973 | *FOXO3* (Forkhead Box O3) | c.1143_1144insG;p.Leu382Alafs*3 | rs758436116 | het | LP | No |
| | 6:26087507 | *HFE* (Homeostatic Iron Regulator) | c.67C>T;p.Arg23Cys | rs761203501 | het | VUS | No |
| | 12:6555481 | *IFFO1* (Intermediate Filament Family Orphan 1) | c.538_549del;p.Ser180_Thr183del | rs756975788 | het | VUS | No |

| | | | | | | | |
|---|---|---|---|---|---|---|---|
| 10 | 5:42718493 | *GHR* (Growth Hormone Receptor) | c.986A>G;p.His329Arg | rs775435127 | het | VUS | No |
| | 6:42684814 | *UBR2* (Ubiquitin Protein Ligase E3 Component N-Recognin 2) | c.4796A>C;p.Lys1599Thr | rs1044454747 | het | VUS | No |
| | 7:100211095 | *STAG3* (Stromal Antigen 3) | c.3323T>C;p.Leu1108Pro | ND | het | VUS | No |
| | 10:119827427 | *INPP5F* (Inositol Polyphosphate-5-Phosphatase F) | c.3046G>T;p.Val1016Phe | rs150134182 | het | VUS | No |
| | 11:94396457 | *GPR83* (G Protein-Coupled Receptor 83) | c.455A>G;p.Tyr152Cys | rs1234084993 | het | VUS | No |
| | 15:89325562 | *POLG* (DNA Polymerase Gamma, Catalytic Subunit) | c.1837C>T;p.His613Tyr | rs147407423 | het | VUS | Ptosis, mypopathy and severe cerebellar atrophy (2) |
| | 19:2249369 | *AMH* (Anti-Mullerian Hormone) | c.37C>G;p.Leu13Val | rs754607106 | het | VUS | No |
| | 20:23048319 | *THBD* (Thrombomodulin) | c.1186C>A;p.Pro396Thr | ND | het | VUS | No |
| | 20:47649093 | *NCOA3* (Nuclear Receptor Coactivator 3) | c.3645_3651+5del | rs770158269 | het | LP | No |
| 11 | 5:42718663 | *GHR* (Growth Hormone Receptor) | c.1156C>T;p.Arg386Cys | rs34853905 | het | VUS | No |
| | 7:144399832 | *NOBOX* (NOBOX Oogenesis Homeobox) | c.1079G>A;p.Arg360Gln | rs199538689 | het | LB | No |
| | 11:108335849 | *ATM* (ATM Serine/Threonine Kinase) | c.8156G>A;p.Arg2719His | rs55982963 | het | VUS | Susceptibility to breast cancer (3) |
| | 17:58218727 | *MKS1* (MKS Transition Zone Complex Subunit 1) | c.83T>C;p.Val28Ala | rs201957874 | het | VUS | No |
| | 17:58212983 | *MKS1* (MKS Transition Zone Complex Subunit 1) | c.857A>G;p.Asp286Gly | rs151023718 | het | VUS | Bardet-Biedl (4) |
| | 17:746992 | *GEMIN4* (Gem Nuclear Organelle Associated Protein 4) | c.1050_1051del;p.Asp350Glufs*28 | rs758240351 | het | VUS | No |
| 12 | 1:54886909 | *DHCR24* (24-Dehydrocholesterol Reductase) | c.211G>T;p.Val71Leu | ND | het | VUS | No |
| | 2:169493760 | *BBS5* (Bardet-Biedl Syndrome 5) | c.542T>C;p.Phe181Ser | rs758508869 | het | VUS | No |
| | 5:42718990 | *GHR* (Growth Hormone Receptor) | c.1483C>A;p.Pro495Thr | rs6183 | het | P | Susceptibility to lung cancer (5) |
| | 20:62837104 | *COL9A3* (Collagen Type IX Alpha 3 Chain) | c.1625C>T;p.Ala542Val | rs753247678 | het | VUS | No |
| | 22:21683186 | *PPIL2* (Peptidylprolyl Isomerase Like 2) | c.482C>T;p.Pro161Leu | rs765685353 | het | VUS | No |

B, benign; Het, heterozygous; Hom, homozygous; LB, likely benign; LP, likely pathogenic; ND, not determined; P, pathogenic; G6PDH, glucose-6-phosphate dehydrogenase; VUS, variant of unknown significance. For each gene, sequence information is based on: ADCY7 (NM_001114.5), AMH (NM_000479.5), ARVCF (NM_001670.3), ATM (NM_000051.4), ATR (NM_001184.4), BBS5 (NM_152384.3), CDH1 (NM_004360.5), CEBPB (NM_005194.4), COL9A3 (NM_001853.4), DHCR24 (NM_014762.4 ), EXO1 (NM_130398.4), FOXO3 (NM_001455.4), G6PD (NM_001360016.2), GEMIN4 (NM_015721.3), GHR (NM_000163.5), GPR83 (NM_016540.4), GRIN2C (NM_000835.6), HFE (NM_000410.4), IFFO1 (NM_001193457.2), IL6ST (NM_002184.4), INPP5F (NM_014937.4), ITIH3 (NM_002217.4), KYAT3 (NM_001008661.3), MKS1 (NM_017777.4), MTRR (NM_002454.3), NBN (NM_002485.5), NCOA3 (NM_181659.3), NF1 (NM_001042492.3), NOBOX (NM_001080413.3), POLG (NM_002693.3), POLM (NM_013284.4), PPIL2 (NM_014337.4), ROS1 (NM_001378902.1), SARDH (NM_001134707.2), STAG3 (NM_001282717.2), THBD (NM_000361.3), UBR2 (NM_001363705.2), VPS18 (NM_020857.3), ZMIZ2 (NM_031449.4)

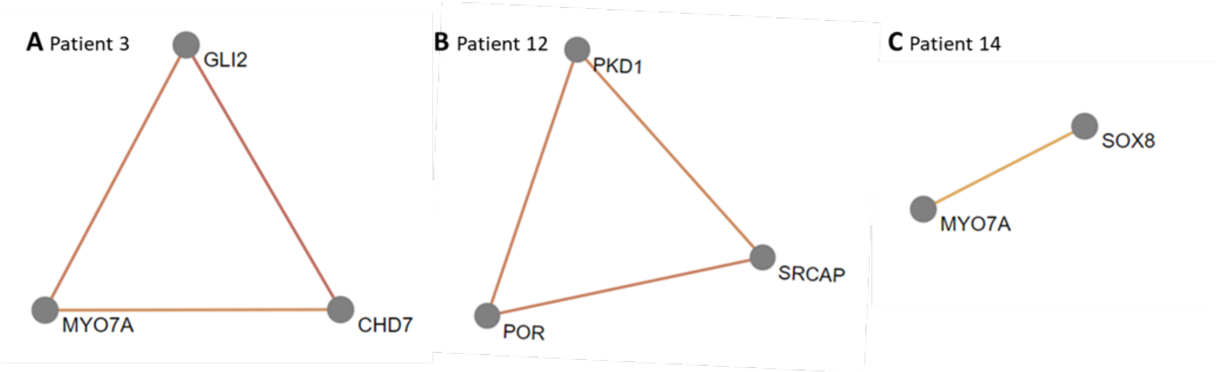


Supplementary figure 1. Potential oligogenic interaction networks of DSD- and *NR5A1*-related genes identified in specific DSD individuals harbouring the *NR5A1*/SF-1 p.Gly146Ala variant. Networks were identified for patients 3, 12 and 14 respectively. To search for potential oligogenic disease networks, the Oligogenic Resource for Variant AnaLysis (ORVAL, https://orval.ibsquare.be/) was used. Nodes represent genes and edges connect two genes only, if between them there is at least one candidate disease-causing variant combination predicted by VarCoPP. The colour of the edge represents the pathogenicity score for that pair of genes. This score is represented in a colour range from brown (higher pathogenicity score) to yellow (lower pathogenicity score).
